# Protection against omicron severe disease 0-7 months after BNT162b2 booster

**DOI:** 10.1101/2022.05.04.22274647

**Authors:** Ofra Amir, Yair Goldberg, Micha Mandel, Yinon M. Bar-On, Omri Bodenheimer, Laurence Freedman, Sharon Alroy-Preis, Nachman Ash, Amit Huppert, Ron Milo

## Abstract

Following a rise in cases due to the delta variant and evidence of waning immunity after 2 doses of the BNT162b2 vaccine, Israel began administering a third BNT162b2 dose (booster) in July 2021. Recent studies showed that the 3rd dose provides a much lower protection against infection with the omicron variant compared to the delta variant and that this protection wanes quickly. In this study, we used data from Israel to estimate the protection of the 3rd dose against severe disease up to 7 months from receiving the booster dose. The analysis shows that protection conferred by the 3rd dose against omicron did not wane over a 7-month period and that a 4th dose further increased protection, with a severe disease rate approximately 3-fold lower than in the 3-dose cohorts.

## Introduction

Following a rise in cases due to the delta variant and evidence of waning immunity after 2 doses of the BNT162b2 vaccine,^1^ Israel began administering a third BNT162b2 dose (booster) in July 2021. The 3rd dose was very effective against both confirmed infections and severe disease caused by the Delta variant.^2^ However, recent studies showed that the 3rd dose provides a much lower protection against infection with the omicron variant and that this protection wanes quickly.^3^

A “fresh” 3rd dose was shown to be effective against severe disease caused by the omicron variant.^4^ However, it is not yet known how long this protection lasts. A study from the UK estimated 85% vaccine effectiveness for individuals 65 years of age or older 15 weeks after receiving the 3rd dose.^5^ Here, we report on the protection against severe disease with the omicron variant in individuals aged 60 or older up to 7 months after receiving a 3rd BNT162b2 dose.

## Methods

### Description of the Data

The Ministry of Health (MOH) in Israel collects all COVID-19 related variables in a central database. These include data on all PCR and antigen tests and results, vaccination dates and type (almost all received the Pfizer-BioNTech vaccine), daily clinical status of all COVID-19 hospitalized patients, and COVID-19 related deaths. Specifically, the data used for conducting this study included vaccination dates, PCR and state-regulated rapid antigen tests (dates and results), hospital admission dates (if relevant), clinical severity status (severe illness or death), and demographic variables such as age, sex, and demographic group (General Jewish, Arab, ultra-Orthodox Jewish).

The data for the study was retrieved on March 26, 2022. Severe illness due to Covid-19 during 14 days following a confirmed infection (defined using the NIH definition^6^ as a resting respiratory rate of more than 30 breaths per minute, an oxygen saturation of less than 94% while breathing ambient air, or a ratio of partial pressure of arterial oxygen to fraction of inspired oxygen of less than 300). Thus, the study period was set from January 16, 2022, to March 12, 2022, to allow for 14 days of follow-up from infection to severe disease. Those who died from COVID-19 during the first 14 days after confirmed infection were also counted as severe disease cases in our analysis. Surveillance of COVID-19-associated hospitalizations is continuously performed by the MOH. Data from all hospitals are updated daily, and often twice a day. In accordance with national guidelines, healthcare providers report all hospitalizations and deaths among individuals with laboratory-confirmed SARS-CoV-2 infection. Quality assurance of data was performed extensively over the course of the pandemic. The data are monitored daily by the MOH, and are continuously used for public health decision-making.

### Study design and population

We conducted an observational study during Israel’s fifth wave, which was omicron-dominant,^7^ with both BA.1 and BA.2 sublineage present (initially BA.1 dominated, later BA.2 dominated). The study included persons who were 60 years of age or older, who received their 2nd dose at least four months before the end of the study, who were not infected by SARS-CoV-2 before the study period, had available data regarding sex and demographic sector, had not stayed abroad during the whole study period, and had not been vaccinated with a vaccine different from BNT162b2 before the study period (see Figure 1).

**Figure 1.**
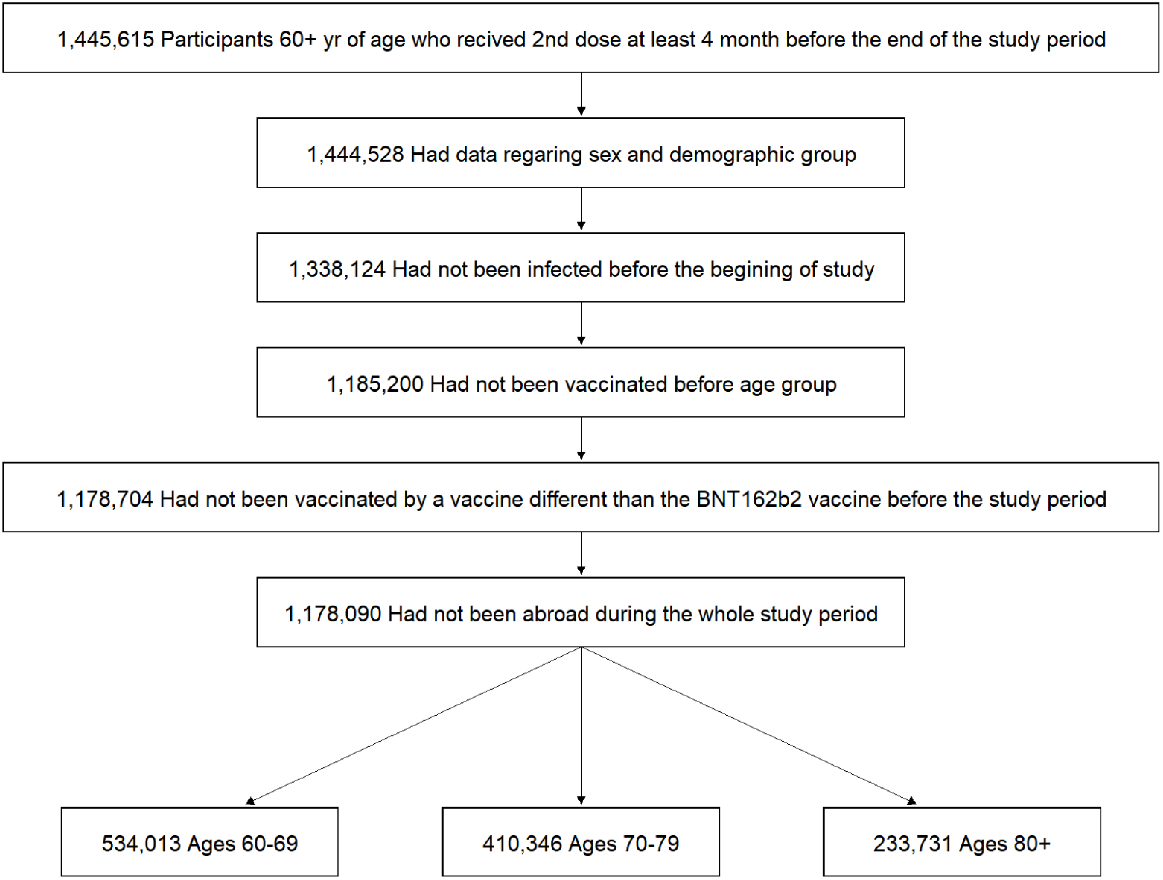
Study population. The participants in the study included persons who were 60 years of age or older, who received 2nd dose at least four months before the end of the study, who were not infected by SARS-CoV-2 before the study period, had available data regarding sex and demographic sector, had not stayed abroad during the whole study period, and had not been vaccinated with a vaccine different from BNT162b2 before the study period.

### Statistical analysis

We estimated the adjusted rates of severe disease in nine cohorts: people who received only 2 doses of the vaccine and for whom at least 4 months had passed since receiving the second dose, people who received a 3rd vaccine dose (divided into 7 cohorts based on the number of months that passed since the administration of that dose), and people who recently received a 4th vaccine dose. Figure 2 shows the vaccination dynamics during the study period, and Table 1 reports basic descriptive statistics for the different cohorts.

**Figure 2.**
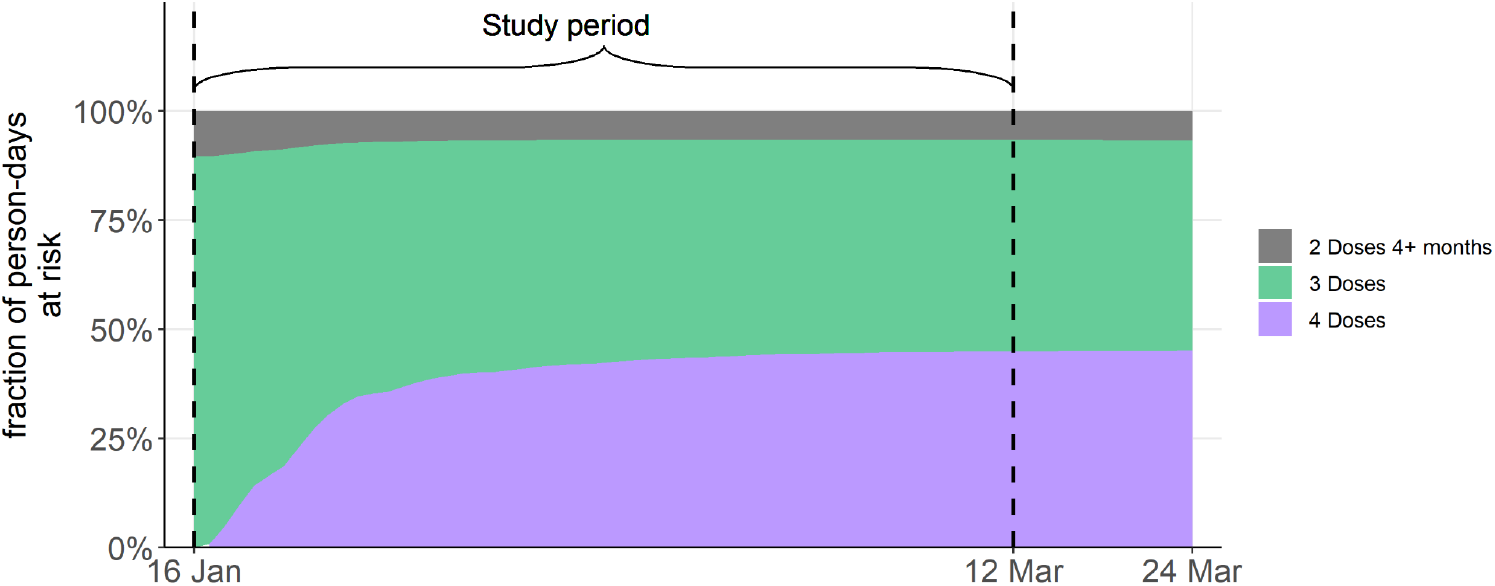
Vaccination dynamics of people aged 60 or above during the study period. The dashed vertical lines represent the study period, between January 16, 2022, and March 12, 2022.

A Poisson regression model was used to estimate the adjusted rate of severe disease per 100,000 risk days. The model included covariates adjusting for age category (60-69, 70-79, 80+), gender, sector (Arab, General Jewish, Ultra-Orthodox), and epidemiological week. We further conducted two sensitivity analyses. First, we allowed for severe illness up to 21 days following a confirmed infection. Second, we examined the adjusted rates of severe disease by age category (60-69, 70-79, and 80+).

## Results and summary

The results of the Poisson regression are summarized in Table 2 and in Figure 3. The protection conferred by the 3rd dose against omicron did not show signs of waning over a 7-month period, with rates of severe disease at approximately 4 per 100,000 risk days in all 3-dose cohorts. This rate was approximately 3-fold lower than in the 2-dose cohort. The 4th dose further increased protection, with a severe disease rate approximately 3-fold lower than in the 3-dose cohorts. Both sensitivity analyses yielded estimates of the rate ratios between the groups that were similar to those obtained in the main analysis.

**Figure 3.**
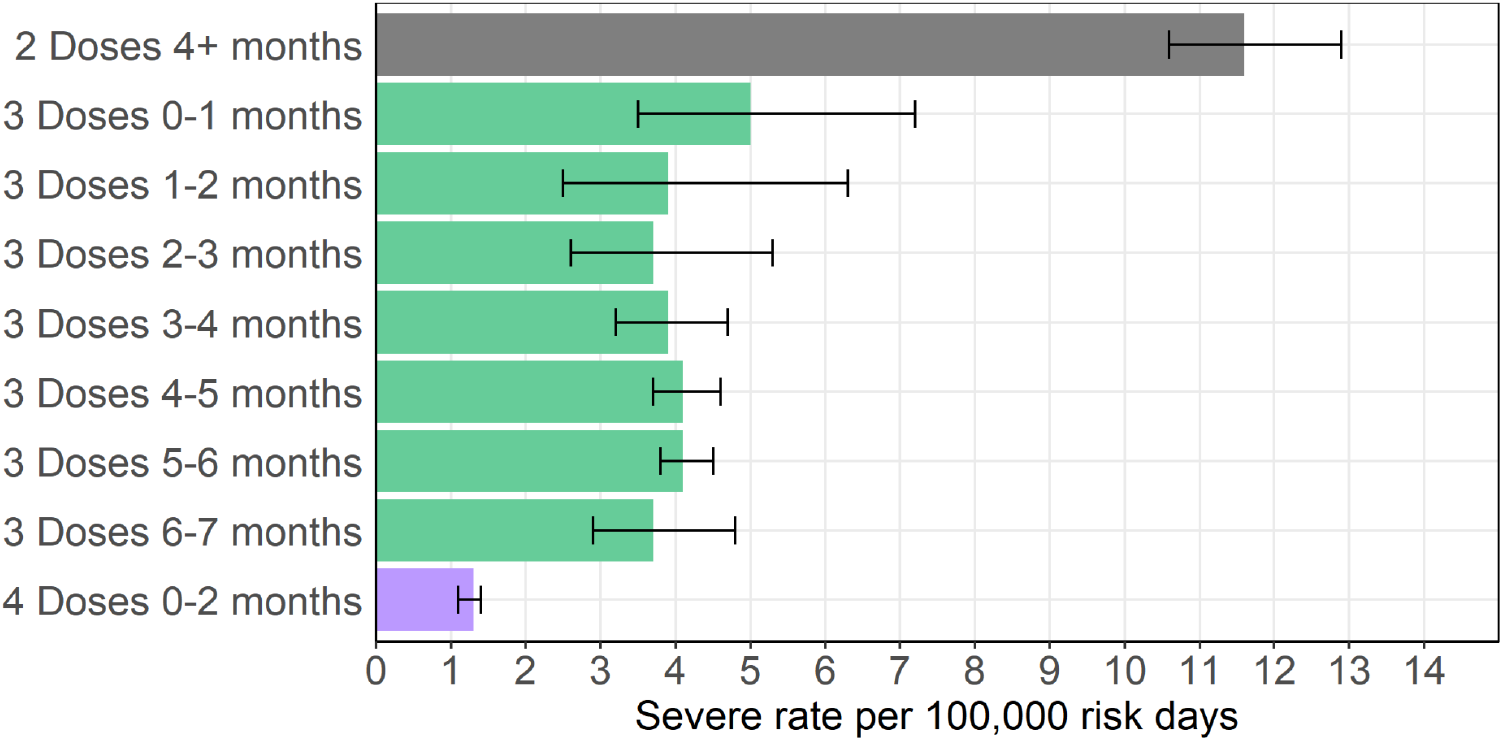
Adjusted rates of severe illness per 100,000 risk days obtained from Poisson regression analysis for the study period January 16, 2022, to March 12, 2022, adjusted for age category (60-69, 70-79, 80+), gender, sector, and exposure (based on the epidemiological week).

Recent studies have shown that the protection of a 3rd BNT162b2 dose against confirmed omicron infections is substantially lower than against the Delta variant, and wanes quickly. Our results show that the effectiveness of the 3rd dose in preventing severe disease does not appear to wane over a 7-month period, and that a 4th dose provides additional protection.

### Ethics statement

The study was approved by the Institutional Review Board of the Sheba Medical Center.

## Data Availability

The individual-level data used in this study cannot be publicly shared even if anonymized due to privacy restrictions.

## Competing interests statement

All authors declare no competing interests.

---

**Table S1.** Demographic and clinical characteristics of the different cohorts. Risk days and infections are calculated for the study period January 16 to March 12, 2022. Severe disease cases were defined as severe illness occurring within 14 days of an infection confirmed in the study period. Rates are per 100,000 person-days. The table presents the proportion of person-days at risk (for the confirmed infection analysis) instead of the number of individuals, as people can move between cohorts.

| Group |  | Total | Female | Male | 60-69 | 70-79 | 80+ | General Jewish | Ultra-Orthodox Jewish | Arabs |
| --- | --- | --- | --- | --- | --- | --- | --- | --- | --- | --- |
| 2 Doses 4+ months<br>Person-days at risk<br>= 3,858,236 | % person days at risk | 100% | 60.80% | 39.20% | 53.50% | 28.00% | 18.50% | 63.20% | 5.70% | 31.10% |
|  | # severe Covid-19 (rate) | 469<br>(12.2) | 236<br>(10.1) | 233<br>(15.4) | 100<br>(4.8) | 142<br>(13.1) | 227<br>(31.9) | 281<br>(11.5) | 18<br>(8.2) | 170<br>(14.2) |
| 3 Doses 0-1 months<br>Person-days at risk<br>=446,008 | % person days at risk | 100% | 59.80% | 40.20% | 47.20% | 30.00% | 22.80% | 77.80% | 5.10% | 17.10% |
|  | # severe Covid-19 (rate) | 29<br>(6.5) | 12<br>(4.5) | 17<br>(9.5) | 5<br>(2.4) | 7<br>(5.2) | 17<br>(16.7) | 22<br>(6.3) | 2<br>(8.8) | 5<br>(6.5) |
| 3 Doses 1-2 months<br>Person-days at risk<br>= 524,213 | % person days at risk | 100% | 60.40% | 39.60% | 51.00% | 29.70% | 19.30% | 77.70% | 4.90% | 17.40% |
|  | # severe Covid-19 (rate) | 18<br>(3.4) | 11<br>(3.5) | 7<br>(3.4) | 3<br>(1.1) | 5<br>(3.2) | 10<br>(9.9) | 12<br>(2.9) | 1<br>(3.9) | 5<br>(5.5) |
| 3 Doses 2-3 months<br>Person-days at risk | % person days at risk | 100% | 61.70% | 38.30% | 51.70% | 29.10% | 19.20% | 76.90% | 4.60% | 18.50% |
| = 683,876 | # severe Covid-19<br>(rate) | 30<br>(4.4) | 16<br>(3.8) | 14<br>(5.3) | 4<br>(1.1) | 8<br>(4) | 18<br>(13.7) | 25<br>(4.8) | 1<br>(3.2) | 4<br>(3.2) |
| 3 Doses 3-4<br>months<br>Person-days at risk<br>= 2,286,002 | % person days at<br>risk | 100% | 59.60% | 40.40% | 57.00% | 27.40% | 15.60% | 78.10% | 4.20% | 17.70% |
|  | # severe Covid-19<br>(rate) | 104<br>(4.5) | 45<br>(3.3) | 59<br>(6.4) | 17<br>(1.3) | 29<br>(4.6) | 58<br>(16.3) | 76<br>(4.3) | 4<br>(4.2) | 24<br>(5.9) |
| 3 Doses 4-5<br>months<br>Person-days at risk<br>= 7,262,548 | % person days at<br>risk | 100% | 57.00% | 43.00% | 56.40% | 28.80% | 14.80% | 81.60% | 4.40% | 13.90% |
|  | # severe Covid-19<br>(rate) | 353<br>(4.9) | 155<br>(3.7) | 198<br>(6.3) | 62<br>(1.5) | 111<br>(5.3) | 180<br>(16.7) | 270<br>(4.6) | 19<br>(5.9) | 64<br>(6.3) |
| 3 Doses 5-6<br>months<br>Person-days at risk<br>=13,018,739 | % person days at<br>risk | 100% | 54.70% | 45.30% | 51.80% | 32.40% | 15.80% | 84.60% | 4.80% | 10.60% |
|  | # severe Covid-19<br>(rate) | 561<br>(4.3) | 220<br>(3.1) | 341<br>(5.8) | 89<br>(1.3) | 171<br>(4.1) | 301<br>(14.7) | 458<br>(4.2) | 28<br>(4.5) | 75<br>(5.4) |
| 3 Doses 6-7<br>months<br>Person-days at risk<br>= 5,942,383 | % person days at<br>risk | 100% | 53.60% | 46.40% | 49.40% | 34.40% | 16.20% | 85.60% | 5.00% | 9.50% |
|  | # severe Covid-19<br>(rate) | 75<br>(1.3) | 21<br>(0.7) | 54<br>(2) | 13<br>(0.4) | 24<br>(1.2) | 38<br>(3.9) | 49<br>(1) | 4<br>(1.4) | 22<br>(3.9) |
| 4 Doses 0-2<br>months<br>Person-days at risk<br>= 20,924,765 | % person days at<br>risk | 100% | 52.60% | 47.40% | 32.00% | 41.70% | 26.30% | 93.90% | 2.70% | 3.50% |
|  | # severe Covid-19<br>(rate) | 280<br>(1.3) | 113<br>(1) | 167<br>(1.7) | 28<br>(0.4) | 83<br>(1) | 169<br>(3.1) | 252<br>(1.3) | 6<br>(1.1) | 22<br>(3) |

**Table 2.** Results of the Poisson regression analysis for severe disease; Adjusted rates of severe disease and adjusted rate ratios compared to the 2nd dose cohort.

| Group<br>(days from<br>vaccination) | Severe cases<br>(person-days at risk) | Adjusted Rate of<br>severe disease per<br>100,000 person-days at<br>risk [95% CI] | Adjusted Rate Ratio<br>relative to 2nd dose<br>cohort [95% CI] |
| --- | --- | --- | --- |
| 2nd dose<br>(4+ months) | 469 (3,858,236) | 11.6 [10.6, 12.9] | Ref |
| 3rd dose<br>(0-1 months) | 29 (446,008) | 5 [3.5, 7.2] | 2.3 [1.6, 3.4] |
| 3rd dose<br>(1-2 months) | 18 (524,213) | 3.9 [2.5, 6.3] | 2.9 [1.8, 4.7] |
| 3rd dose<br>(2-3 months) | 30 (683,876) | 3.7 [2.6, 5.3] | 3.1 [2.2, 4.6] |
| 3rd dose<br>(3-4 months) | 104 (2,286,002) | 3.9 [3.2, 4.7] | 3.0 [2.4, 3.7] |
| 3rd dose<br>(4-5 months) | 353 (7,262,548) | 4.1 [3.7, 4.6] | 2.8 [2.5, 3.3] |
| 3rd dose<br>(5-6 months) | 561 (13,018,739) | 4.1 [3.8, 4.5] | 2.8 [2.5, 3.2] |
| 3rd dose<br>(6-7 months) | 75 (5,942,383) | 3.7 [2.9, 4.8] | 3.1 [2.4, 4.1] |
| 4th dose<br>(0-2 months) | 280 (20,924,765) | 1.3 [1.1, 1.4] | 9.2 [7.9, 10.7] |

## Notes

### Competing Interest Statement

The authors have declared no competing interest.

### Funding Statement

This study did not receive any funding

### Author Declarations

The study was approved by the Institutional Review Board of the Sheba Medical Center.

